# Single-nucleus multiomics in brains from Hispanic individuals reveal *APOE-ε4*-driven disruption of focal adhesion signaling in the presence of cerebrovascular pathology

**DOI:** 10.1101/2025.05.21.25328040

**Authors:** Elanur Yilmaz, Kevin W. Chen, Elif Öykü Cakir, Prabesh Bhattarai, Minghua Liu, Benjamin L. Ciener, Min Qiao, Rafael Lantigua, Martin Medrano, Diones Rivera, Angel L. Piriz, Philip De Jager, Francesca Bartolini, Yiyi Ma, Andrew F. Teich, Annie J. Lee, Dolly Reyes-Dumeyer, Badri N. Vardarajan, Richard Mayeux, Caghan Kizil

**Affiliations:** Department of Neurology, Columbia University Irving Medical Center, New York, NY, USA; Taub Institute for Research on Alzheimer’s Disease and the Aging Brain, New York, NY, USA; Center for Translational and Computational Neuroimmunology, Department of Neurology, Columbia University Irving Medical Center, New York, NY 10032, USA; Department of Medicine, Vagelos College of Physicians and Surgeons, Columbia University Irving Medical Center, New York, New York, USA; School of Medicine, Pontificia Universidad Catolica Madre y Maestra Santiago, Dominican Republic; Department of Neurology, CEDIMAT, Plaza de la Salud, Santo Domingo, Dominican Republic; School of Medicine, Universidad Pedro Henriquez Urena (UNPHU), Santo Domingo, Dominican Republic; Gertrude H. Sergievsky Center, Vagelos College of Physicians and Surgeons, Columbia University Irving Medical Center, New York, NY, USA; Department of Pathology and Cell Biology, Columbia University Irving Medical Center; New York, NY 10032, USA; Department of Psychiatry, Columbia University Irving Medical Center, New York, NY 10032, USA; Department of Epidemiology, Mailman School of Public Health, Columbia University Irving Medical Center, New York, NY 10032, USA

**Author notes:** Correspondence to C.K. Contributed equally (Badri N. Vardarajan, Richard Mayeux, Caghan Kizil).

## Abstract

The apolipoprotein E ε4 allele (*APOE-ε4*) is the strongest genetic risk factor for late-onset Alzheimer’s disease (AD), yet its molecular impact on cerebrovascular biology remains inconclusive, particularly in underrepresented populations with elevated vascular burden. Individuals from Hispanic ancestry experience disproportionately high rates of cerebrovascular pathology, offering a unique opportunity to investigate the mechanisms of cerebrovascular pathology in AD. Here, we performed single-nucleus RNA sequencing (snSeq) on 413,175 nuclei from 52 postmortem Hispanic brains to determine *APOE-ε4*-associated cell type specific transcriptomic changes in a population with elevated cerebrovascular risk. We identified a conserved molecular signature marked by dysregulated extracellular matrix deposition and focal adhesion signaling in astrocytes. These findings were replicated in the non-Hispanic ROSMAP cohort (n = 424) snSeq. Findings were validated in isogenic human iPSC-derived astrocytes, humanized *APOE* targeted replacement mouse brains, and post-mortem human brains at protein and chromatin accessibility level. Our data suggest that *APOE-ε4* astrocytes adopt a hyper-adhesive, mechanically rigid phenotype that may exacerbate cerebrovascular pathology.

## Introduction

Alzheimer’s disease (AD), the most prevalent form of dementia, is characterized by amyloid-beta plaques and tau neurofibrillary tangles^1,2^. Yet, growing evidence highlights the critical role of cerebrovascular pathology in AD that may contribute to the cognitive decline and disease progression^3–8^. Cerebrovascular pathologies, include large and small ischemic infarcts and hemorrhages that cause blood-brain barrier (BBB) disruption^9–13^. Therefore, a deeper understanding of the molecular and cellular mechanisms linking cerebrovascular pathology to AD is essential to identify pathways that drive neurodegeneration. A key approach to achieving this is the study of populations where cerebrovascular burden is particularly pronounced. Individuals of Hispanic ancestry exhibit a disproportionately high prevalence of cardiovascular disease and cerebrovascular risk factors, such as hypertension, diabetes, and metabolic syndrome^14,15^. Autopsy studies further indicate that Hispanic individuals with dementia are more likely to present with significant cerebrovascular lesions compared to non-Hispanic whites, emphasizing a stronger cerebrovascular component in this population^16^.

Apolipoprotein E *ε4* allele (*APOE-ε4*), the strongest genetic risk factor for late-onset AD^17–19^. While *APOE-ε4* is known to influence both amyloid deposition and neurodegeneration, emerging evidence suggests that its impact on cerebrovascular health may be comparably consequential^18,20,21^. Understanding how *APOE-ε4* interacts with cerebrovascular dysfunction in various populations, where the burden is high, may provide critical insights into the molecular drivers of AD. Astrocyte-specific removal of *APOE-ε4* ameliorates cerebrovascular dysfunction despite increased amyloid angiopathy^21^ and *APOE-ε4* directly impairs BBB integrity *in vivo*^20^.

We chose single nucleus RNA-sequencing (snRNA-seq) in brains to identify *APOE-ε4*–mediated mechanisms in AD to enable high-resolution, cell-type–specific transcriptomic profiling of complex brain tissue. snRNA-seq preserves the cellular heterogeneity of the brain, allowing us to pinpoint how APOE-ε4 influences distinct populations. Therefore, we performed single nucleus sequencing from 52 available Hispanic brain tissues in New York Brain Bank (NYBB) collected through Washington Heights and Inwood Community Aging project (WHICAP)^22–24^, Estudio Familiar de Influencia Genetica en Alzheimer (EFIGA)^25–28^ and National Institute on Aging Alzheimer’s Disease Family Based Study (NIA-ADFBS)^28,29^. We provide a resource for Hispanic brain multiome, and altered molecular mechanisms biologically related to *APOE* genotypes. We used stringent gene filtering, pathway prioritization, cross-validation in the Religious Order Study/Memory and Aging Project (ROSMAP) single nucleus database. Biological validation in iPSC-derived human cells, humanized *APOE-ε4* expressing mouse brains and post-mortem human brains (**Fig. 1**), showed that focal adhesion signalling is the most significantly affected pathway and is largely related to astrocyte-vasculature interactions. By revealing astrocyte-related cell adhesion deficits, our study not only provides a resource in Hispanic population, but also uncovers a gliovascular mechanism with universal relevance to vascular contribution to AD.

**Figure 1.**
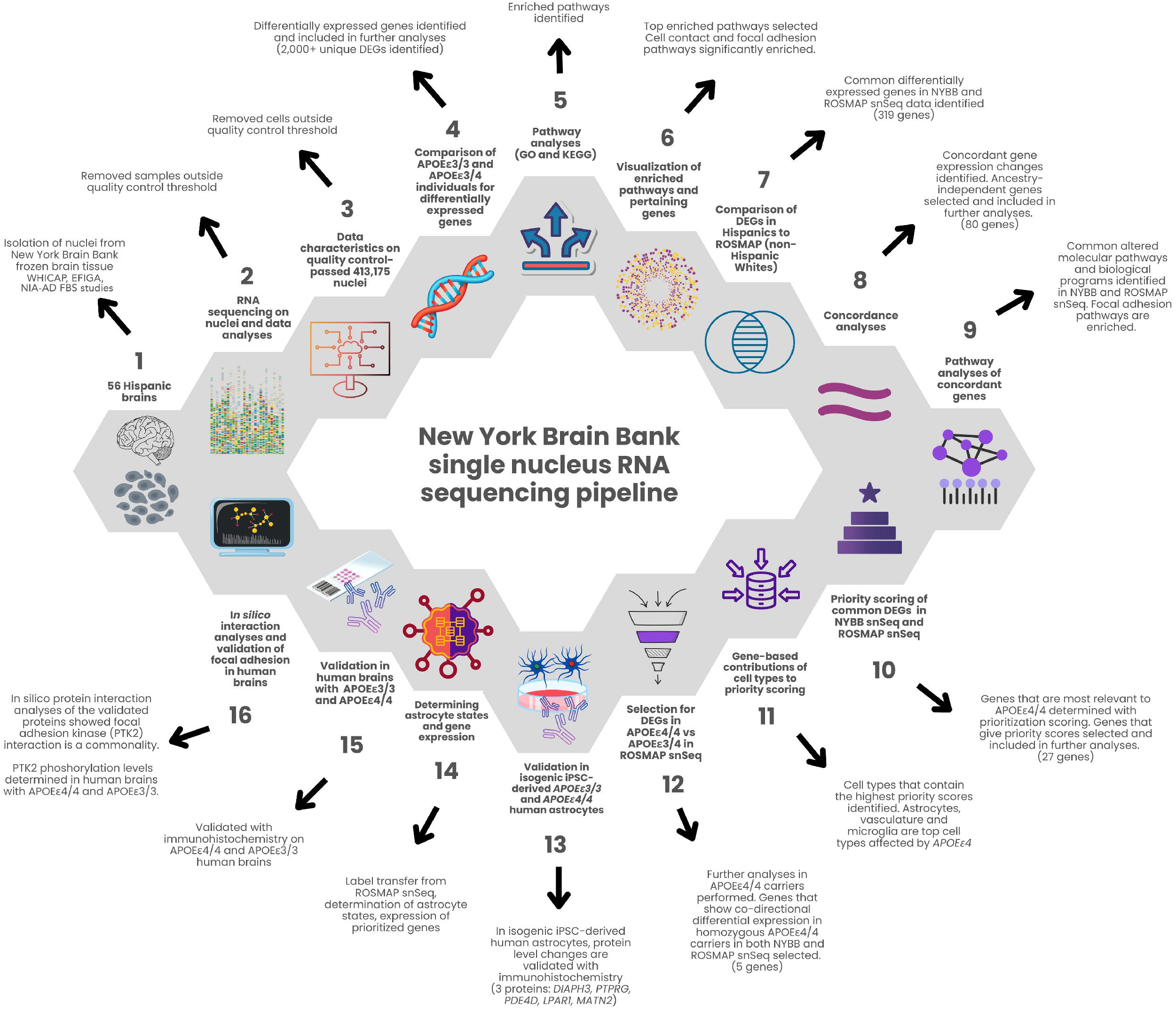
Single-nucleus RNA sequencing and analysis pipeline. (1) Nuclei were isolated from 52 genetically confirmed Hispanic donor brains. (2) Multiome (RNA + ATAC) libraries were generated and sequenced; raw reads were aligned to GRCh38 with Cell Ranger. (3) Samples and nuclei failing quality thresholds (fewer than 200 genes detected or >20% mitochondrial reads) were removed, yielding 413,175 high-quality nuclei. (4) Differential expression analysis comparing APOE-ε3/4 versus APOE-ε3/3 genotypes identified over 2,000 unique DEGs. (5) Gene Ontology and KEGG enrichment analyses revealed altered biological pathways. (6) Cell-contact and focal adhesion pathways emerged as top-ranked and were selected for further investigation. (7) Cross-cohort comparison with ROSMAP snRNA-seq data from non-Hispanic White donors identified 319 shared DEGs. (8) Filtering for concordant regulation across both cohorts yielded 80 ancestry-independent DEGs. (9) Enrichment of extracellular matrix organization and focal adhesion signaling was confirmed on this gene set. (10) A composite priority score (– log₁₀ p-value, log₂ fold change, and percentage of expressing cells) was calculated for each DEG. (11) The contribution of each major cell type to total priority was quantified, with astrocytes, vascular cells, and microglia contributing most. (12) APOE-ε4 allele-dose comparisons (ε4/4 vs. ε3/4) in both NYBB and ROSMAP datasets refined the candidate list to five genes. (13) Immunofluorescence in isogenic APOE-ε3/3 and ε4/4 iPSC-derived astrocytes validated protein-level changes for DIAPH3, PTPRG, PDE4D, LPAR1, and MATN2. (14) Label transfer of ROSMAP-defined astrocyte subtypes onto NYBB nuclei resolved ten distinct astrocyte states and mapped prioritized gene expression. (15) Immunohistochemistry of postmortem APOE-ε3/3 and ε4/4 human brain sections confirmed reduced focal adhesion kinase and AKT phosphorylation in astrocyte endfeet. (16) In silico protein–protein interaction analysis (STRING v12, Cytoscape) revealed a conserved focal adhesion network centered on PTK2 (FAK), AKT, and cytoskeletal regulators.

## Results

### Single-nucleus transcriptomics in Hispanic brains identifies *APOE-ε4*-associated differentially expressed genes

We performed single-nucleus RNA sequencing (snRNA-seq) and quality control analyses of 413,175 nuclei from available 52 postmortem frozen brain samples from Hispanic individuals (38 individuals with Alzheimer’s disease, 5 individuals with mild cognitive impairment, 3 individuals with Parkinson’s disease, 1 individual with Lewy body disease, and 5 control individuals based on clinical intake diagnosis). We used all available frozen brain tissues from people with Hispanic ancestry in New York Bran Bank. The genotypes were as follows: 20 individuals with *APOE-ε3/4* (15 females, 5 males), 14 individuals with *APOE-ε3/3* (9 females, 5 males), 12 individuals with *APOE-ε2/3* (8 females, 4 males), 4 individuals with *APOE-ε2/4* (3 females, 1 male), and 2 individuals *APOE-ε4/4* (both females) (**Extended Data Table 1**).

As per our data generation and quality control pipeline (**Fig. 2a**), RNA counts per nucleus were normally distributed (**Fig. 2b**). Gene and read counts were consistent across genotypes, but *APOE-ε4/4* tissue samples had slightly lower values due to limited sample availability (**Fig. 2c**). Number and type of cells sequenced for every individual (n=52) were consistent, and oligodendrocytes, neurons, microglia, and astrocytes are the most abundant cell types sequenced (**Fig. 2d,e**). UMAP clustering (**Fig. 2f**) and marker gene analyses (**Fig. 2g**) identified astrocytes, microglia, neurons, oligodendrocytes, oligodendrocyte progenitors, endothelial cells, and pericytes (**Fig. 2h-j**). Most nuclei came from *APOE-ε3/3* and *APOE-ε3/4* individuals (69.7%; 44.1% AD and 25.6% non-AD) (**Fig. 2k, l; Extended Data Tables 2, 3**). Cell type proportions and sequencing complexity were similar across genotypes (**Fig. 2m, n; Extended Data Table 4**). Expression levels of two housekeeping genes (*ACTB* and *GAPDH*) were consistent within samples (**Fig. 2o**). In our cohort, the number of *APOE-ε4* alleles per individual positively correlates with Braak stage (r = 0.41), cerebral amyloid angiopathy (r = 0.13) and arteriolosclerosis (r = 0.22), while negatively correlates with age (r = –0.22). In contrast, the number of *APOE-ε3* alleles per individual is negatively correlated with Braak stage (r = −0.22), cerebral amyloid angiopathy (r = −0.19) and arteriolosclerosis (r = −0.22) and shows positive correlation with age (r = 0.22) (**Fig. 2p**). Additionally, individuals with increasing number of *APOE-ε4* alleles tend to show higher levels of CAA and arteriolosclerosis; in contrast, *APOE-ε3* alleles are more frequently observed in older individuals with milder pathology (**Fig. 2q**). Therefore, comparing *APOE-ε4* carriers to *APOE-ε4* non*-*carriers could allow isolating the biological consequences of *APOE-ε4*. We continued the analyses with *APOE-ε3/3* and *APOE-ε3/4* samples, which yielded the most nuclei.

**Figure 2.**
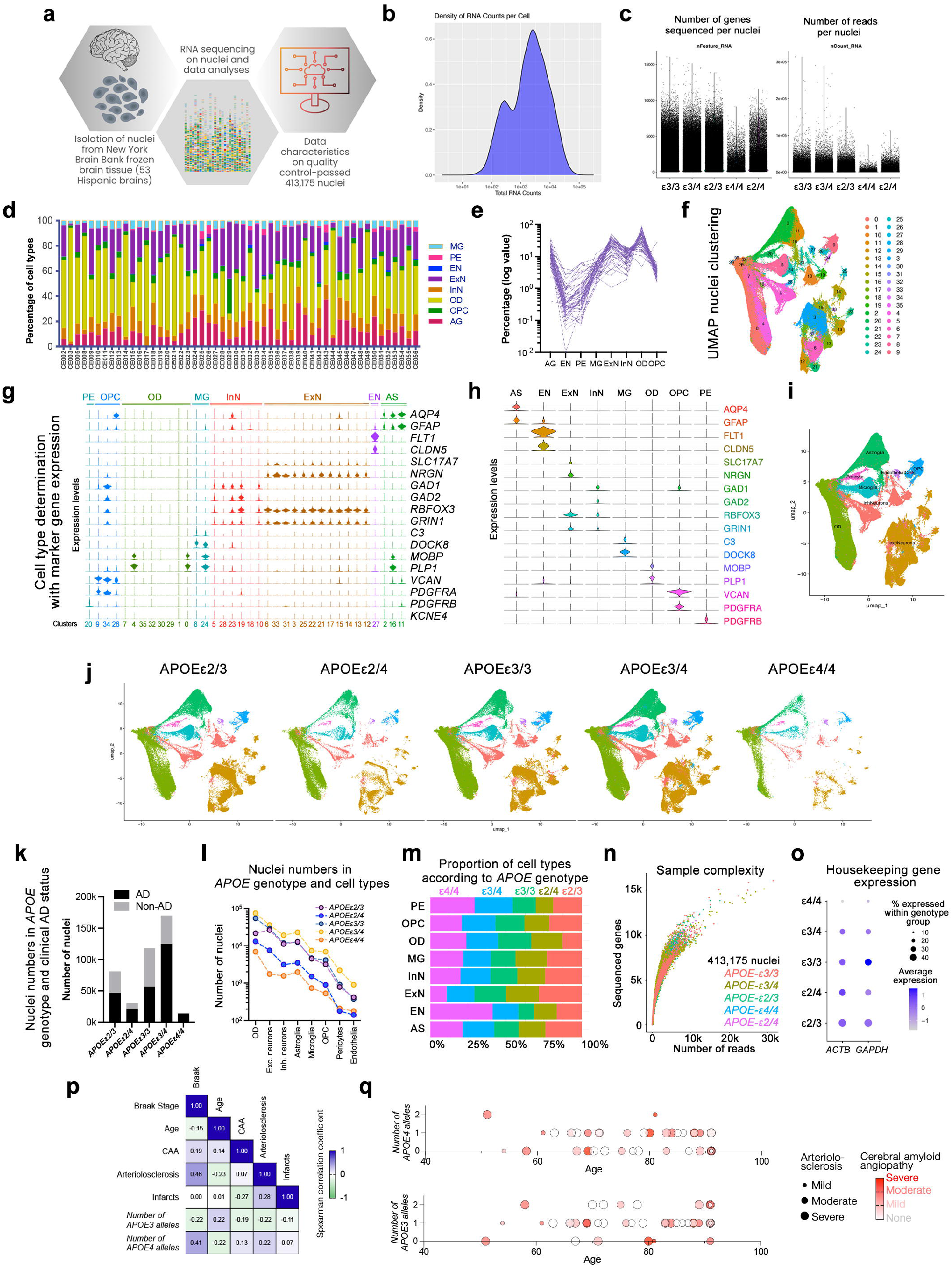
Single-nucleus RNA sequencing of 52 Hispanic human brains reveals *APOE* genotype-specific cellular landscapes. **(a)** Schematic overview of the study part design. Nuclei were isolated from frozen postmortem brain tissue from the New York Brain Bank (52 Hispanic brains) and subjected to single-nucleus RNA sequencing (snRNA-seq), followed by data quality control and computational analyses. A total of 413,175 high-quality nuclei were included. **(b)** Density plot showing the distribution of total RNA counts per nucleus across the dataset. The bimodal pattern reflects expected variability in RNA capture among nuclei. **(c)** Scatter plot showing the number of genes detected per nucleus and number of reads per nucleus, stratified by APOE genotype (ε2/3, ε2/4, ε3/3, ε3/4, ε4/4). Despite some variability, gene detection was generally comparable across genotypes. Read depth distributions were consistent across genotypes, indicating that APOE status did not introduce systematic bias in sequencing coverage. **(d)** Color-coded percent representation of sequenced cells for 52 individuals sequenced in this study. **(e)** Percent representation of every cell type sequenced in every individual (connected dotted lines represent one individual). **(f)** UMAP plot showing transcriptomic clustering of nuclei. Each color represents a distinct cluster identified through unsupervised analysis, capturing transcriptional heterogeneity across brain cell types. **(g)** Marker gene expression across clusters used to assign major brain cell identities. Rows show canonical marker genes, and columns correspond to clusters, with dot size and color indicating expression level and proportion of expressing cells, respectively. **(h)** Combined cell types and their distinctive markers shown in violin plots. **(i)** Resulting cell type classification on combined UMAP. **(i)** Annotated UMAP with cluster identities manually labeled based on marker gene expression. Major brain cell types include astrocytes (AS), microglia (MG), oligodendrocytes (OD), neurons (ExN, InN), endothelial cells (EN), pericytes (PE), and oligodendrocyte progenitor cells (OPC). **(j)** UMAP projections stratified by APOE genotype. Distribution of nuclei from each APOE genotype group (ε2/3, ε2/4, ε3/3, ε3/4, ε4/4) visualized separately to assess genotype-specific clustering or compositional shifts. **(k)** Number of nuclei recovered per APOE genotype, further stratified by clinical Alzheimer’s disease (AD) status. AD and non-AD nuclei counts are shown in black and gray, respectively. **(l)** Line plot showing the number of nuclei per cell type across APOE genotypes. Each point represents a genotype, highlighting genotype-specific differences in cell type abundance. **(m)** Stacked bar plot showing the proportion of each cell type within APOE genotypes. Cell type representation varies with APOE genotype, with notable enrichment of endothelial cells in APOE-ε4/4. **(n)** Sample complexity analysis showing the number of genes detected per nucleus as a function of sequencing depth (number of reads), color-coded by APOE genotype. **(o)** Dot plot displaying expression of housekeeping genes (ACTB and GAPDH) across APOE genotypes. Dot size indicates percentage of cells expressing the gene, and color reflects average expression, confirming consistency across groups for normalization. **(p)** Spearman correlation matrix of clinical and pathological features including Braak stage, age, cerebral amyloid angiopathy (CAA), arteriolosclerosis, infarcts, and number of *APOE-ε3* and *APOE-ε4* alleles. The strength and direction of associations are represented by the color scale, where blue indicates positive correlations and green indicates negative correlations. **(q)** Dot plots showing the relationship between age and the number of *APOE-ε4* (top) and *APOE-ε3* (bottom) alleles. Each dot represents an individual, with dot size indicating arteriolosclerosis severity (larger size = more severe), and dot color indicating cerebral amyloid angiopathy (CAA) severity from none (white) to severe (red).

We identified differentially expressed genes (DEGs) by comparing *APOE-ε3/4* to *APOE-ε3/3* nuclei. Most DEGs appeared in only one or two cell types (**Fig. 3a,c; Extended Data Table 5**). KEGG enrichment analysis revealed significant changes in focal adhesion, MAPK signaling, oxidative phosphorylation, Alzheimer’s disease, and axon guidance pathways (**Fig. 3d, e**). Focal adhesion signaling was altered mainly in astrocytes and vascular cells (**Fig. 3f**). Many extracellular matrix (ECM) components and adhesion regulators, including *FN1, COL4A2, COL6A2, LAMA2, LAMA3, IGF1, TJP1, PXN, ZYX*, were differentially expressed (*FN1, COL6A2, LAMA2, PXN, ZYX* upregulated and *LAMA3, IGF1, TJP1* downregulated in astrocytes) (**Fig. 3f**), which we validated in *APOE-ε3/3* and *APOE-ε3/4* brains with immunostainings (**Fig. 3g**). Gene ontology (GO) enrichment supported ECM and adhesion pathway alterations, including integrin binding and basement membrane interactions (**Fig. 3h, i; Extended Data Fig. 1**). Other affected pathways, such as adherens junctions, insulin and neurotrophin signaling, also showed disruptions in adhesion-related genes (**Extended Data Fig. 2**). These included downregulated *PTPRG* (a phosphatase that negatively regulates focal adhesion kinase)^30,31^*, RAC1* (a small GTPase that regulates cytoskeletal tension and mechanotransduction signalling)^32^*, PDE4D* (phosphodiesterase that promotes cAMP-PKA signalling and altering adhesion stability)^33,34^, and integrin-activating neurotrophin genes. These results suggested a prominent *APOE-ε4*-induced focal adhesion disruption, primarily in astrocytes and vascular cells **(Fig. 3; Extended Data Figs. 1 and 2).**

**Figure 3.**
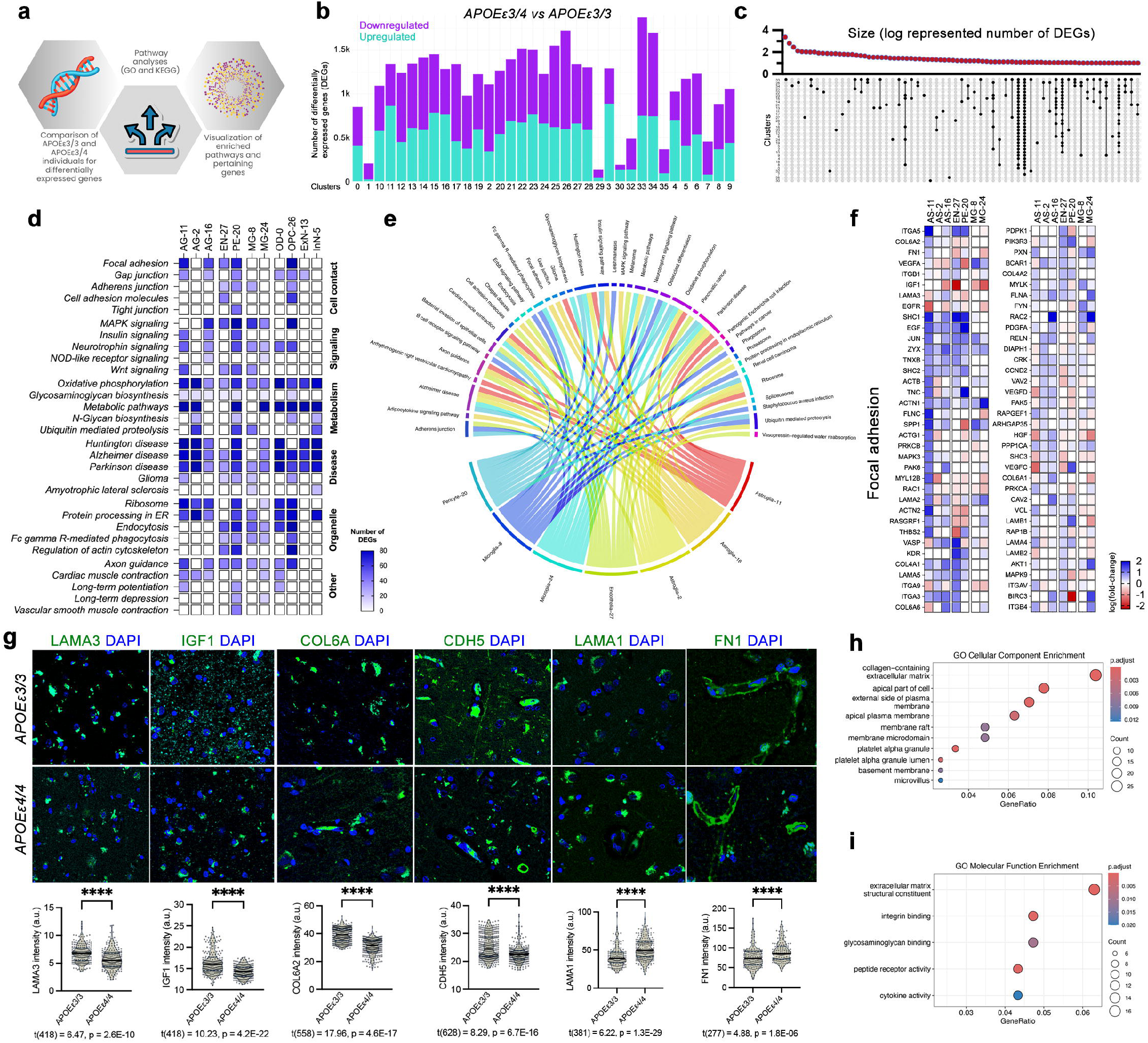
Differential gene expression and pathway enrichment analysis in *APOE-ε4* carriers reveals focal adhesion dysregulation. (a) Schematic overview of the analytical workflow. Differentially expressed genes (DEGs) were identified by comparing brains from *APOE-ε3/4* and *APOE-ε3/3* genotypes, followed by enrichment analysis to identify significantly altered pathways. **(b)** Bar graph showing the number of upregulated (purple) and downregulated (cyan) DEGs across all cell clusters in *APOE-ε3/4* versus *APOE-ε3/3* brains, indicating widespread transcriptomic alterations. **(c)** DEG distribution across clusters, displayed on a log-transformed scale. Each dot corresponds to the number of DEGs per cluster. DEGs in multiple cell clusters are linked with a vertical line. **(d)** Heatmap of the number of DEGs in top altered pathways. **(e)** Chord diagram linking DEGs to the top enriched pathways, illustrating shared genes involved in multiple signaling categories. **(f)** Heatmap of focal adhesion-related DEGs across astrocyte, microglia and vascular cell clusters. **(g)** Validation of protein expression changes in postmortem brain sections from *APOE-ε3/3* and *APOE-ε4/4* individuals. Representative immunofluorescence images and quantification of LAMA3, IGF1, COL6A, CDH5, LAMA1 and FN1 expression levels. Statistical significance was determined using two-tailed unpaired parametric t-test. t value, degrees of freedom (in parenthesis), and p values are given under the graphs. Data are presented as mean ± s.e.m. ****p < 0.0001. At least 3 individuals used for every group. **(h)** GO Cellular Component enrichment of DEGs in *APOE-ε3/4* versus *APOE-ε3/3* individuals in NYBB snSeq. **(i)** GO Molecular Function enrichment of DEGs in *APOE-ε3/4* versus *APOE-ε3/3* individuals in NYBB snSeq.

### Cross-cohort validation confirms *APOE-ε4*-dependent alterations in focal adhesion signaling

We replicated these findings using snSeq data from the ROSMAP cohort^35–37^. We tested the generalizability of the findings from our Hispanic cohort that bears a higher cerebrovascular comorbidity load^15,16,27,28^, in a predominantly non-Hispanic white ROSMAP cohort^38^ (**Fig. 4; Extended Data Table 6**). We identified 319 common DEGs (21.4% of all DEGs in ROSMAP) between the two datasets (**Extended Data Table 7**), with 296 showing co-directional changes in at least one corresponding cell type (**Fig. 4b**). GO term and Reactome enrichment analyses revealed consistent involvement of ECM components, basement membrane structures, and integrin signaling pathways (**Fig. 4c**). These findings suggest a shared *APOE-ε4*-associated gliovascular molecular signature that is a genotype-driven mechanism.

**Figure 4.**
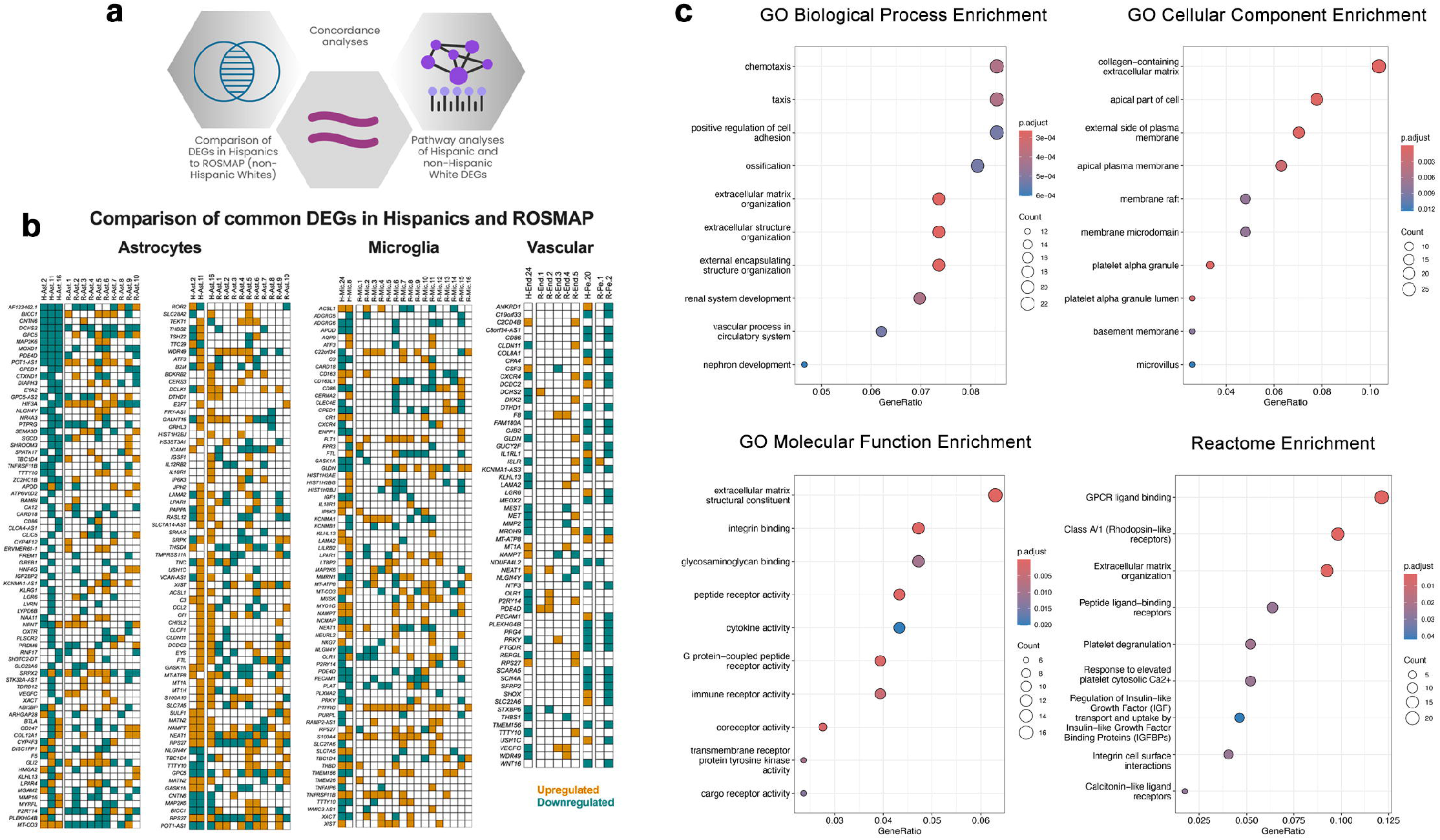
Cross-cohort analysis of differentially expressed genes in Hispanic and non-Hispanic AD brains reveals conserved alterations in extracellular matrix and focal adhesion signaling. **(a)** Schematic overview of the concordance analysis workflow. Differential gene expression (DEG) profiles were compared between astrocytes, microglia, and vascular cells from Hispanic AD brains (NYBB dataset) and non-Hispanic White AD brains (ROSMAP cohort). Overlapping DEGs were subjected to gene ontology and pathway enrichment to determine conserved and population-specific biological processes. **(b)** Comparison heatmap of common DEGs in astrocytes, microglia, and vascular cells across the two datasets. Each row represents a gene; columns represent cell clusters in NYBB (left) and ROSMAP (right). Orange tiles indicate upregulated genes, and teal tiles indicate downregulated genes. Shared genes between datasets highlight conserved molecular alterations across populations, particularly in pathways related to ECM remodeling, focal adhesion, and inflammation. **(c)** Gene Ontology (GO) and Reactome pathway enrichment analyses performed on shared DEGs in astrocytes from the NYBB cohort. Enrichment significance was calculated using the hypergeometric test with Benjamini-Hochberg correction for multiple comparisons. Adjusted p-values (p.adjust) are shown as color gradients in the bubble plots. Bubble size reflects the number of genes enriched in each term, and the x-axis represents the gene ratio (number of genes in the term relative to all DEGs analyzed).

### Gene prioritization highlights focal adhesion-related genes

We prioritized the 296 DEGs using a scoring method based on three non-correlating parameters: p-values, fold change, and the percentage of cells expressing the gene, capturing significant, highly expressed cell-specific genes with robust statistical support, as we documented before^39,40^ (**Extended Data Fig. 3; Extended Data Table 8**). Astrocytes and vascular cells (endothelia and pericytes) held most high-priority scores (23.2% and 29.9% of the total scores obtained for all cell clusters, respectively; **Fig. 5a**). We shortlisted 80 genes with scores above 0.1 threshold in at least one cell type (**Fig. 5b**). Top candidates included mitochondrial genes (*MT-CO3, MT-ATP8*), ECM components and regulators (*TMEM156, SGCD, JPH2, MATN2, MMP16*), adhesion modulators (*PTPRG, PTPRQ, SULF1, PDE4D, PLXNA2, DLCK1, PAPPA*), and cell communication-modifying receptors (*LPAR1, LPAR2, GPC3, GPC5, KDR*). We mapped the distribution of scores of every gene by cell type (**Fig. 5c**). Five genes, *CPA4, THBS2, ZNF114, SEMA6A-AS2, and DTHD1*, showed >90% of their scores in a single cell type. Sixteen genes had a dominant cell type (>50%) and a secondary contributor (>20%). *MATN2, FTL*, *CFI*, *TSHZ2*, *ATP6V0D2*, *FXYD3* (dominant in astrocytes), *GALNT15*, *APOD*, *EYS, EYAZ* (dominant in oligodendrocytes or progenitors), *MGAM2, SLA2*, *ADGRG6, CYP26B1*, *GPC3* (dominant in neurons), and NLRP12 (dominant in endothelia) (**Fig. 5c**). Other 59 genes showed broad distribution, suggesting widespread and pleiotropic *APOE-ε4* effects.

**Figure 5.**
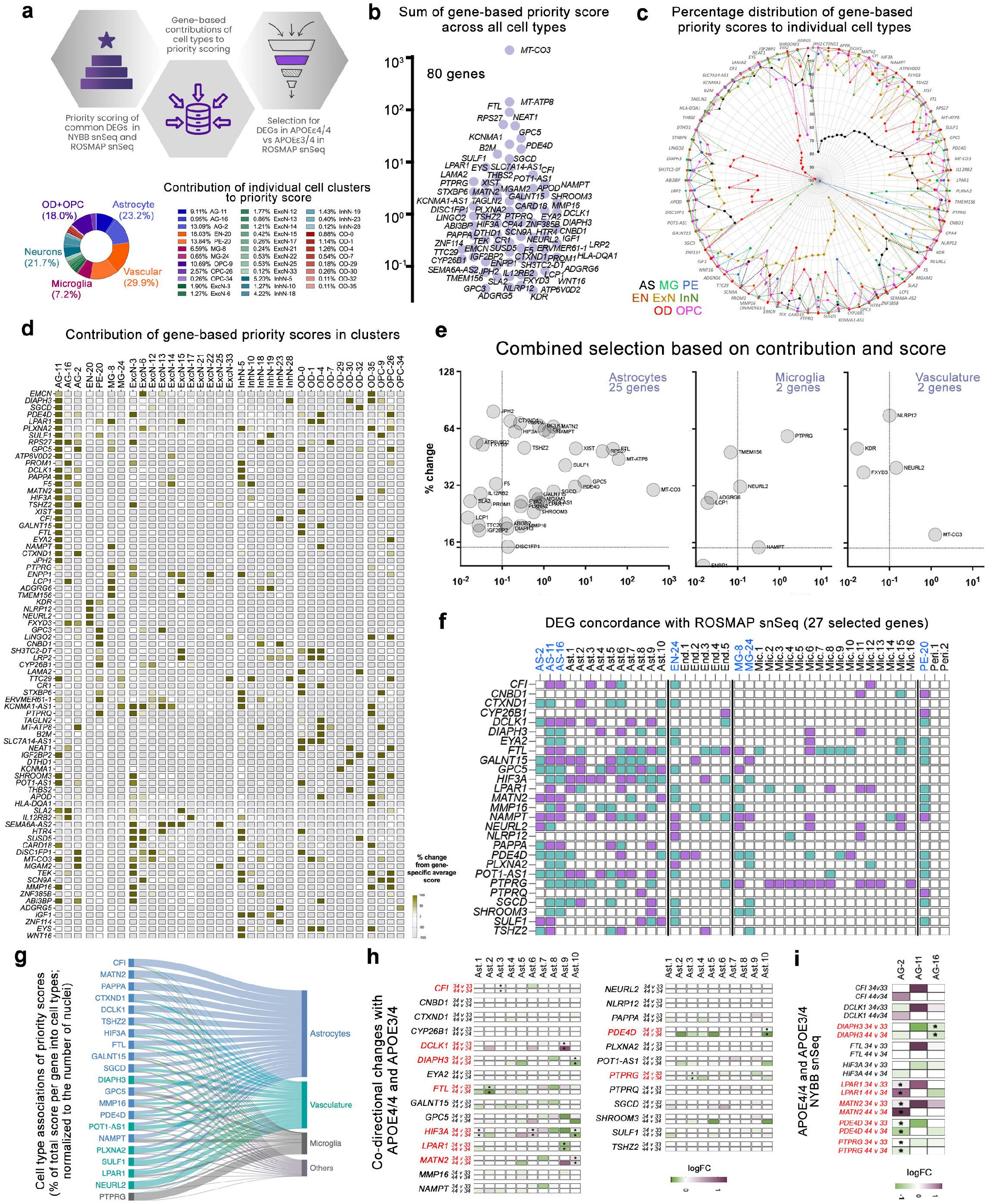
Prioritization of differentially expressed genes in *APOE-ε4* astrocytes highlights focal adhesion changes across multiethnic cohorts. (a) Schematic of the prioritization strategy. Common DEGs between Hispanic and non-Hispanic White (ROSMAP) snSeq datasets were assigned cell-type-specific priority scores based on three non-correlated parameters: log fold change, statistical significance, and percentage of expressing cells. The contribution of each cell type to the total priority score is visualized in the pie chart. Astrocytes (23.2%) and vascular cells (29.9%) were the dominant contributors. **(b)** Log-scale plot of the summed priority scores for 80 selected DEGs across all cell types. Mitochondrial genes (e.g., MT-CO3, MT-ATP8) and ECM-regulatory genes (FTL, PDE4D, LPAR1) rank among the highest. **(c)** Circular barplot of the percentage distribution of gene-based priority scores per cell type. Each gene’s dominant cell-type specificity is indicated, showing that many top candidates are shared between astrocytes, vasculature, and oligodendrocyte lineages. **(d)** Heatmap displaying the contribution of each cell cluster to individual gene priority scores. Yellow shading indicates clusters with >50% of a gene’s priority score, suggesting cluster-specific expression relevance. **(e)** Combined selection of 27 final genes based on score magnitude and cell-type preference. Genes with the highest scores and specific enrichment in astrocytes (left), microglia (middle), and vasculature (right) were prioritized. **(f)** Heatmap showing DEG direction concordance between NYBB and ROSMAP snSeq datasets for the 27 prioritized genes. Green and purple boxes indicate directionally consistent up- or downregulation across at least one cluster in both datasets. **(g)** Sankey diagram illustrating the cell-type associations of priority scores for the 22 genes concordant in NYBB and ROSMAP snSeq data. Most genes were linked to astrocytes, with additional contributions from microglia and vascular cells. **(h)** Table showing co-directional gene expression changes between *APOE-ε4/4* vs. *APOE-ε3/4* (in NYBB snSeq) and *APOE-ε3/4* vs. *APOE-ε3/3* (in ROSMAP snSeq), confirming allele-dose effects. Genes highlighted in red (e.g., *CFI, DCLK1, FTL, LPAR1, MATN2, PDE4D, PTPRG*) show progressive regulation with APOE4 dosage. **(i)** Final validation of 27 genes in NYBB snSeq data comparing *APOE-ε4/4* vs. *APOE-ε3/4*. Log2 fold changes are shown for astrocyte clusters; genes with consistent directional change (highlighted in red) were considered robust *APOE-ε4*-responsive candidates. Gene priority scores were computed using a composite score integrating log2 fold change, adjusted p-values (Benjamini-Hochberg corrected), and percentage of expressing cells per cluster. Concordance across datasets and allele-dose consistency was assessed in binary mode (upregulated/downregulated) and by directionality of log2 fold change. Enrichment significance in earlier stages of analysis (not shown here) was calculated using the hypergeometric test with multiple testing correction.

To further prioritize the genes, we examined whether specific genes had disproportionately high priority scores in individual clusters compared to the average score for that cell type. This approach identifies the genes that may be particularly relevant to a specific physiological or functional state within a cell lineage (**Extended Data Table 8**). Many genes showed stronger scores in specific cell clusters, implying that their expression was more concentrated in particular subset compared to the rest of those cell types. We identified 43 genes in astrocytes, 15 in vascular cells (we considered endothelia and pericytes together as vascular cells), 12 in microglia, 33 in excitatory neurons, 34 in inhibitory neurons, 35 in oligodendrocytes, and 27 in oligodendrocyte progenitor cells (**Fig. 5d**) that were differentially expressed in *APOE-ε4* carriers. We selected astrocyte- and vascular-enriched genes using score (>0.1) and cluster specificity (>15%) cutoffs, which we chose to balance combinatorial stringency and to capture the genes with both substantial overall priority and meaningful specificity within a cell cluster. This yielded in identification of 27 high-confidence genes (**Fig. 5e**). Two genes, *PTPRG* and *NEURL2*, also appeared in microglia. DEG comparisons across NYBB and ROSMAP cohorts revealed 22 genes with concordant differential expression in at least one cell cluster of the corresponding cell type in both cohorts (**Fig. 5f**). Most genes (20/22) scored highest in astrocytes (**Fig. 5g**), which warranted further focus on downstream analyses.

Because of the availability of *APOE-ε4/4* carriers in the ROSMAP single cell data, we tested DEGs in *APOE-ε4/4*, *APOE-ε3/4* and *APOE-ε3/3* individuals. This allowed us to validate an *APOE-ε4* dose-dependent effect of genes identified in the Hispanics (**Fig. 5h; Extended Data Table 9**). Nine genes showed *APOE-ε4* dose-dependent changes. In NYBB astrocyte DEGs (**Extended Data Table 10**). Five genes - *DIAPH3, LPAR1, MATN2, PDE4D,* and *PTPRG*, showed co-directional changes between *APOE-ε4/4* vs *APOE-ε3/4*, and *APOE-ε3/4* vs *APOE-ε3/3* comparisons (**Fig. 5i**). These five genes consistently show *APOE-ε4* dose-dependent expression changes across datasets and cell types, marking them as strong candidates involved in *APOE-ε4*-driven biological alterations.

### Prioritized genes are related to astrocyte states

To investigate whether our top candidates exhibit state-specific patterns, we performed label transfer from the ROSMAP astrocyte reference^35^ onto the Hispanic astrocyte nuclei (**Extended Data Fig. 4a**). This defined ten distinct astrocyte states (Ast-1 through Ast-10) (**Extended Data Fig. 4b**), with state markers reminiscent of the ROSMAP data^35^ (**Extended Data Fig. 4c**). Feature-plot visualization revealed that *PDE4D* and *PTPRG* are broadly expressed across most states, whereas *DIAPH3, MATN2,* and *LPAR1* are confined to specific subpopulations (**Extended Data Fig. 4d**). Astrocyte state-resolved violin plots confirmed that, *MATN2* and *LPAR1* peak in Ast.4/5, which are documented reactive astrocyte states^35^ (**Extended Data Fig. 4e**). These data indicate that *APOE-ε4*-associated focal adhesion genes not only differ between genotypes but also delineate transcriptionally distinct astrocyte states.

### Prioritized genes have differential chromatin modeling

To determine whether prioritized gene expression was influenced by AD status, we stratified all cell nuclei and astrocyte nuclei by both genotype (ε3/ε3 vs. ε3/ε4) and clinical diagnosis (AD vs. non-AD). The expression levels of *PDE4D, DIAPH3, PTPRG,* and *LPAR1* remained consistent within genotype groups regardless of AD status, supporting that these transcriptional changes are genotype-driven and not consequential to AD pathology (**Extended Data Fig. 5**).

To determine whether the transcriptional remodeling we observed could be due to the changes in chromatin state, we analyzed snATAC-seq data from the same Hispanic multiome samples. We found that *APOE-ε3/4* astrocytes and endothelia/pericytes as vascular cells exhibit differential accessibility at prioritized gene loci (**Extended Data Fig. 6**). For example, at the *PDE4D, PTPRG* and *DIAPH3* loci, *APOE-ε3/4* nuclei display reduced accessibility at promoter-proximal and intronic peaks compared with *APOE-ε3/3*, while conversely, *MATN2* show increased accessibility in *APOE-ε3/4* astrocytes (**Extended Data Fig. 6**). These genotype-dependent shifts suggest cell-type-specific epigenetic mechanisms that may underlie transcriptional changes leading to *APOE-ε4*-driven BBB dysregulation.

### *APOE-ε4* affects PTK2/AKT signalling

To validate the changes in the prioritized genes and their relevance to *APOE-ε4* in astrocytes, we performed *in vitro* and postmortem human brain protein-level analyses (**Fig. 6a**). We found protein expression changes of DIAPH3, PDE4D, PTPRG, LPAR1 and MATN2 in *in vitro* cultures of isogenic *APOE-ε3/3* and *APOE-ε4/4* astrocytes using immunofluorescence. DIAPH3, PDE4D, PTPRG were significantly downregulated while LPAR1 and MATN2 were upregulated in *APOE-ε4/4* cells (**Fig. 6b**). Except for LPAR1, these changes were concordant with the gene expression changes in snSeq. To determine the global protein changes in these astrocytes, we performed quantitative proteomics between *APOE-ε4/4* and *APOE-ε3/3* astrocytes and detected protein fragments from 8,664 proteins. We identified 774 proteins as differentially expressed (abs(logFC)>1 and p<0.05) (**Extended Data Table 11**). Pathway analysis of differentially expressed proteins showed altered focal adhesion, ECM-receptor interaction, and actin remodeling pathways (**Fig. 6d**). We built a protein-protein interaction network using the five prioritized genes, *DIAPH3, PDE4D, PTPRG, MATN2* and *LPAR1*. PTK2 (essential component of the focal adhesion kinase) emerged as a central node (**Fig. 6e**).

**Figure 6.**
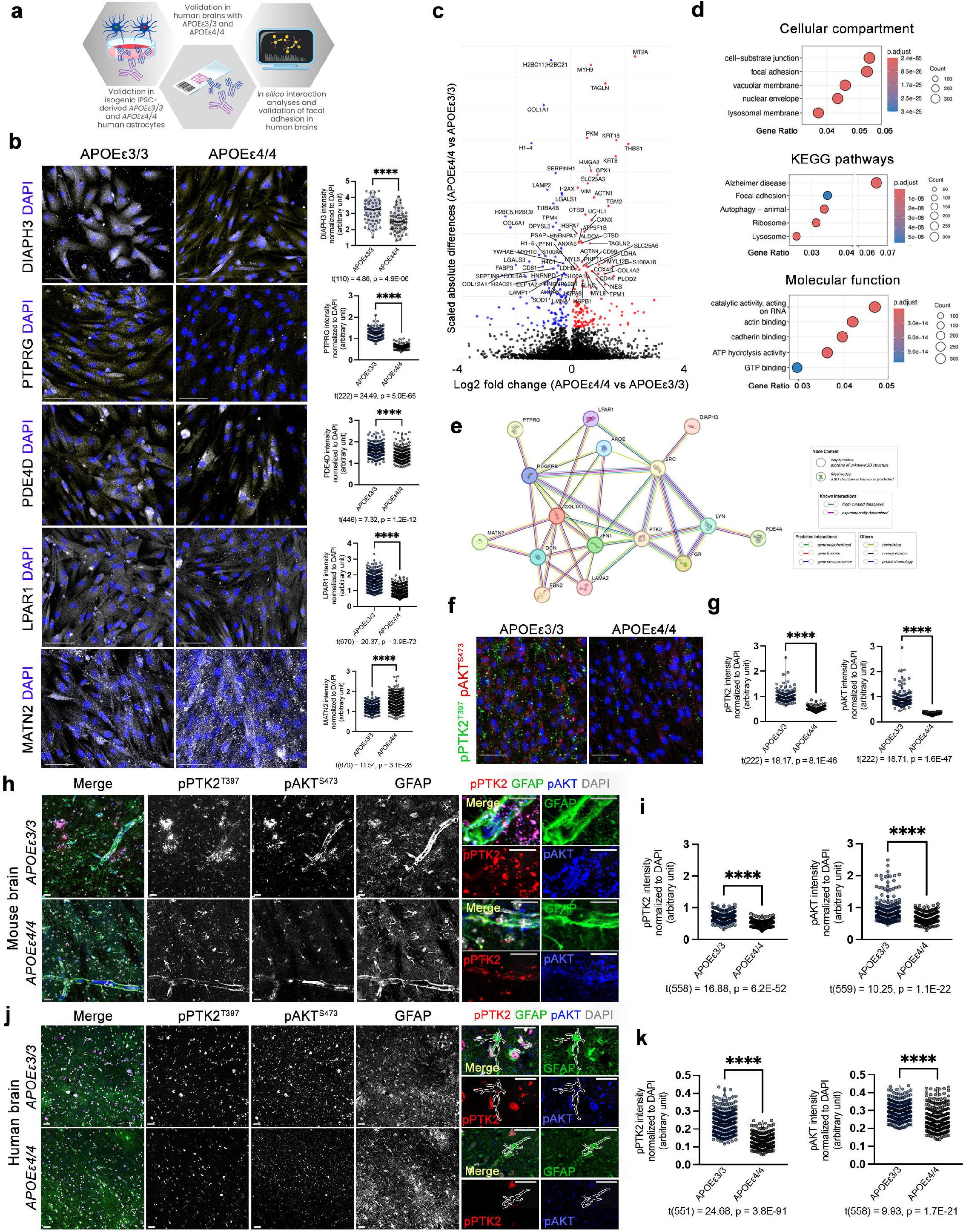
*APOE-ε4* astrocytes exhibit focal adhesion signaling dysregulation marked by reduced PTK2 activation and elevated AKT signaling. (a) Overall validation strategy, which integrates transcriptomic prioritization of focal adhesion-related genes with protein-level validation in postmortem human brain tissue, isogenic iPSC-derived astrocytes, and in silico protein interaction analysis. **(b)** Immunofluorescence staining for DIAPH3, PTPRG, PDE4D, LPAR1 and MATN2 in *APOE-ε3/3* and *APOE-ε4/4* iPSC-derived human astrocytes. Quantification of fluorescence intensity revealed statistically significant reductions for all three proteins, using parametric unpaired two-tailed t-tests, with all data presented as mean ± s.e.m. and significance threshold set at ****p < 0.0001. **(c)** Volcano plot of proteomics. Differentially expressed proteins between *APOE-ε4/4* and *APOE-ε3/3* astrocytes are shown. **(d)** GO and KEGG enrichment analyses of the differentially expressed proteins. Adjusted p-values were calculated using the Benjamini-Hochberg correction method. **(e)** STRING-based protein-protein interaction network based on DIAPH3, PTPRG, PDE4D, LPAR1, and MATN2. PTK2 (FAK) is a central node forming a tightly connected interaction module. **(f)** Immunostaining of isogenic iPSC-derived human astrocytes with *APOE-ε4/4* or *APOE-ε3/3* demonstrates markedly reduced levels of phosphorylated PTK2 (pPTK2^Y397^) and phosphorylated AKT (pAKT^S473^) in *APOE-ε4/4* astrocytes. **(g)** Quantification graphs confirm significant differences between *APOE-ε3/3* and *APOE-ε4/4* for both markers pPTK2 and pAKT. Parametric unpaired two-tailed t-tests. Data presented as mean ± s.e.m. and significance threshold set at ****p < 0.0001. **(h)** Immunostaining for GFAP, pPTK2^Y397^, and pAKT^S473^ on brain sections from targeted replacement *APOE-ε3/3* and *APOE-ε4/4* mouse. Right panels are high magnification of individual fluorescence channels. **(i)** Quantification graphs for (h), confirming *in vitro* results. Parametric unpaired two-tailed t-tests, with all data presented as mean ± s.e.m. and significance threshold set at ****p < 0.0001. At least 3 animals used for every group. **(j)** Immunostaining for GFAP, pPTK2^Y397^, and pAKT^S473^ on postmortem brain sections from *APOE-ε3/3* and *APOE-ε4/4* individuals. Right panels are high magnification of representative outlines astrocytes. **(k)** Quantification graphs for (j), confirming *in vitro* and mouse results. Parametric unpaired two-tailed t-tests, with all data presented as mean ± s.e.m. and significance threshold set at ****p < 0.0001.At least 3 individuals used for every group.

To test whether PTK2 activity and downstream signalling were affected by *APOE-ε4*, we measured phosphorylation of PTK2 at tyrosine-397 (pPTK2^T397^) and AKT at serine-473 (pAKT^S473^). Both were significantly reduced in *APOE-ε4/4* astrocytes (**Fig. 6f, g**). We confirmed reduced focal adhesion signaling in postmortem mouse and human *APOE-ε4/4* brains in comparison to *APOE-ε3/3* (**Fig. 6h-k**). Astrocytes contacting blood vessels and their endfeet showed significantly reduced pPTK2^T397^and pAKT^S473^ levels (**Fig. 6i, k**). These results suggested that *APOE-ε4* may disrupt focal adhesion signaling via impaired PTK2 and AKT activity.

## Discussion

In this study, we conducted single-nucleus RNA sequencing on 52 postmortem brains from individuals of Hispanic ancestry and identified robust *APOE-ε4*-associated molecular signatures, including changes in focal adhesion signaling. Through a multi-step prioritization framework, cross-cohort validation, and functional protein-level assays, we identified a conserved set of genes linked to focal adhesion dynamics in astrocytes. Our results propose focal adhesion signalling as a target molecular mechanism that could be addressed for mitigating AD risk.

We began our study in Hispanic individuals due to their elevated burden of cerebrovascular pathology^14,16,41–43^, which provides a sensitized background for uncovering cerebrovascular mechanisms of neurodegeneration. We generalized our findings by comparing results to the non-Hispanic ROSMAP cohort^35–37,44^. The consistency across both ancestries strengthens the relevance of our findings, supports the presence of generalizable mechanisms of *APOE-ε4*-driven risk, and identify a pathological alteration of focal adhesion dynamics.

Our data indicate that *APOE-ε4* astrocytes adopt a hyperadhesive phenotype at the gliovascular interface. These cells exhibit reduced PTK2 (focal adhesion kinase, FAK) signaling and impaired morphological adaptability, producing a mechanically rigid state. This rigidity may impair neurovascular coupling and astrocyte-mediated clearance, as we have previously seen with functionally validated human *FMNL2* variants that prevent astrocyte endfeet retraction from the vasculature^45^.

PTK2 is a mechanosensitive integrator of cell-ECM signaling^46^. PTK2 initiates focal adhesion turnover through force-mediated release from autoinhibition, allowing activation and coordination with actin cytoskeletal remodeling^47^. In our study, the diminished phosphorylation of PTK2^Y397^ and its downstream effector AKT^S4^^73^ in *APOE-ε4* astrocytes suggests a failure to engage this mechanotransduction cycle. Without remodeling, astrocyte endfeet remain locked in a rigid adhesive state, consistent with prior findings on genetic variant in a filopodial protein-coding gene *FMNL2*^45^, which act together in the filopodia with *DIAPH3*^48^ – one of our prioritized genes. This extends the focal adhesion activity and gliovascular interactions into a disease-relevant context, supporting the hypothesis that gliovascular rigidity in *APOE-ε4* brains may stem from impaired mechanical signaling at the adhesion complex. PTK2 protein levels or subcellular localization are also reduced in *APOE-ε4/4* astrocytes as we determined in our proteomics analyses, further limiting its activation at focal adhesion sites. These converging deficits may result in structurally persistent but functionally weakened adhesions, producing a signaling-deficient, mechanically rigid astrocyte phenotype.

Chronic elevation of ECM blunts focal adhesion signaling through persistent FAK desensitization. First, extracellular signal-regulated kinase 1/2 (ERK1/2) recruited by FAK activates a tyrosine phosphatase that, with PIN1, dephosphorylates PTK2^Y397^, limiting kinase activity under sustained ECM engagement^49^. Second, overloaded STX6–mediated recycling routes divert excess ECM–integrin complexes to lysosomes, depleting surface integrins and further weakening FAK signaling^50^. This is supported by our findings that, STX6 protein levels are elevated with *APOE-ε4/4* in astrocytes (**Extended Data Table 11**), and PIN1 is upregulated in astrocytes (**Extended Data Table 5**). Together, these feedback loops ensure that once ECM activity exceeds an optimal threshold, focal adhesion signaling is dampened.

The gene expression changes we observed further support our hypothesis. *PDE4D* downregulation may reduce cAMP-PKA signaling^51^, which normally tempers adhesion strength via phosphorylation of adhesion proteins^33^. This loss may allow excessive integrin activation without proper turnover. Altered levels of *LPAR1* could also promote RhoA-ROCK–mediated cytoskeletal rearrangements as a compensatory response to reduced FAK signaling^52^, and *MATN2* could further stabilize adhesions by enhancing ECM-integrin engagements^53^ or inflammation^54^. Together, these changes anchor astrocyte endfeet more tightly to the vascular basement membrane, locking them into an adhesive state that lacks the plasticity needed to respond to neurovascular cues. This molecular context converges on a common failure of dynamic adhesion remodeling, a process critical for glymphatic clearance - the astrocyte-mediated removal of interstitial waste^55–58^. Thus, our findings highlight a unifying model of gliovascular rigidity in *APOE-ε4* carrier brains. These findings are supported by several single-nucleus RNA-seq data analyzing AD cell states in the vasculature, in astrocytes, and in other cell types that reported similar alterations^3,35,36,59–64^. The inability of astrocytes to disengage from the vasculature as well as other cell-ECM interactions in vascular niche cells may compromise vascular plasticity, impair fluid transport via the glymphatic system, and amplify the risk of neurodegeneration by exacerbating toxic protein aggregation.

Our findings also point to a potential feedforward mechanisms by which ECM components such as fibronectin and matrilin may exacerbate the vascular contribution to AD. *APOE-ε4* promotes fibronectin accumulation at the BBB, disrupting astrocyte-endothelial signaling and impairing clearance mechanisms^25^. Elevated fibronectin levels, especially in *APOE-ε4* carriers leads to loss of BBB integrity and reduced gliovascular decoupling^25,65^. ECM remodeling and cellular response to cell-cell communication are also altered by neuroinflammatory signals such as TNFα and IL1, which are elevated in AD brain^66^. These cytokines alter ECM composition and link reactive gliosis to BBB dysfunction. This may position inflammation not merely as a consequence, but as an upstream driver of adhesive pathology in the gliovascular niche.

Cross-cohort astrocyte state analysis (**Extended Data Fig. 4**) further refines the gliovascular rigidity model by showing that focal adhesion modulators are unevenly distributed among astrocyte states. The broad expression of *PDE4D* and *PTPRG* suggests they underpin general astrocyte–ECM interactions, whereas state-restricted genes like *DIAPH3, MATN2*, and *LPAR1* are specialized subpopulations poised for dynamic endfeet remodeling. Such heterogeneity could explain why some astrocytes are more susceptible to *APOE-ε4*-induced dysfunction altering adhesion dynamics. Similarly, chromatin accessibility analyses (**Extended Data Fig. 6**) showed that *APOE-ε4* not only changes astrocyte and vascular transcriptomes but can alter chromatin landscapes at focal adhesion loci in a cell-type-specific manner. The concordance between accessibility changes and expression of *PDE4D, DIAPH3, PTPRG, LPAR1* and *MATN2* suggests that *APOE-ε4* exerts its effects, at least in part, through epigenetic remodeling as suggested before^44,67^, potentially by altering recruitment of transcription factors or chromatin remodelers at these regulatory elements.

The results from this work indicate that in the presence of *APOE-ε4,* astrocytes are locked in a dysfunctional, contractile state, impeding their ability to dynamically engage with vascular signals. The resulting loss of gliovascular flexibility contributes to perivascular fibrosis, impaired perfusion, and a heightened vulnerability to neuroinflammatory cues. However, the impact of these molecular disruptions likely extends beyond astrocytes. Endothelial cells, pericytes, microglia and oligodendrocytes also exhibit *APOE-ε4*-associated gene expression changes^3,4,35,36,63,64,68–73^, and focal adhesion dysregulation impairs endothelial barrier integrity or pericyte attachment, contributing to vascular leakage and BBB breakdown. Each of these cell types should be further explored as co-contributors to the pathological adhesion landscape in *APOE-ε4* carriers. The gliovascular alterations described here help explain changes in imaging biomarkers in *APOE-ε4* carriers, such as microvascular lesions, reduced perfusion, and elevated phosphorylated tau protein levels. Future work should assess whether targeting components of this adhesion machinery can restore astrocyte plasticity and BBB integrity in *APOE-ε4* models. These pathways may represent tractable therapeutic targets to delay or prevent gliovascular dysfunction in AD.

## Methods

### Human brain samples

Frozen postmortem human brain tissues from 52 genetically confirmed individuals with Hispanic ancestry (**Extended Data Table 1**) were obtained from the New York Brain Bank (NYBB) at Columbia University. Brains were selected without stratification for clinical Alzheimer’s disease (AD) diagnosis to enable unbiased *APOE* genotype-specific analysis. Sample metadata including genotype, clinical status, age, sex, vascular pathology are detailed in **Extended Data Table 1**. Informed consent was obtained and all procedures were approved by the Institutional Review Board at Columbia University.

### APOE genotyping

DNeasy Blood & Tissue Kit (Qiagen, Cat #69504) was used to purify the genomic DNA by following the manufacturer’s protocol. The exon 4 of *APOE* gene was amplified for the two *APOE*-relevant SNPs (rs7412 and rs429358) by using the described primers^74^ (*forward: AGCCCTTCTCCCCGCCTCCCACTGT and reverse: CTCCGCCACCTGCTCCTTCACCTCG*). PCR reaction for each DNA sample was set up with a 25 µl reaction volume containing 1X Q5 reaction buffer, 0.4 µM each primer, 200 µM dNTP, 0.02 U/µl Q5 DNA Polymerase and 1X GC Enhancer (NEB, Cat #M0491L). 50-150 ng gDNA used for each reaction. Amplification was performed with an initial denaturation at 98°C for 10 min followed by 35 cycles at 98°C for 1 min, 68.5 °C for 1 min, and 72 °C for 1 min, and a final extension at 72 °C for 10 min. The size and quality of the PCR products were visualized in 1% agarose gel and purified by using GeneJET PCR Purification Kit (Thermo Scientific, Cat. #K0702). The purified PCR products were sent to Genewiz (South Plainfield, NJ) for Sanger Sequencing with a designed primer 5’-TCGGAACTGGAGGAACAACTGACC-3’.

### Nuclei isolation and snRNA-seq library preparation

For the nuclei isolation from the frozen human brain tissue, Chromium Nuclei Isolation Kit with RNase Inhibitor (16 rxns, PN-1000494) was used. Up to 50 mg human brain tissue per sample was utilized for nuclei isolation. Every brain sample was weight-measured before the isolation. The manufacturer’s instructions were followed for the nuclei isolation. In short, this step involved tissue dissociation and cleanup with lysis buffer, debris removal and washing steps, and resuspension. After the visualization and determination of the final nuclei concentration, 10X Genomics Chromium Next GEM Single Cell Multiome workflow was used by following the manufacturer’s protocol, which includes transposition, GEM generation and barcoding, post-GEM incubation cleanup, library pre-amplification PCR, single nucleus ATAC library construction, cDNA amplification, gene expression library construction, and sequencing. The multiome RNA-Seq libraries were sequenced on the NovaSeqX in JP Sulzberger Columbia Genome Center. The multiome ATAC-Seq libraries were sequenced on the Element Aviti. The data was analyzed using cellranger arc 2.0.2.

### snRNA-seq data processing and quality control

Raw FASTQ files were aligned to the GRCh38 reference genome using Cell Ranger (10x Genomics). Quality control filtering was performed by removing nuclei with expressing less than 200 genes, genes expressed less than 3 nuclei, and nuclei expressing more than 20% mitochondrial RNA genes out. Integrated Seurat object was analyzed using Seurat (version 4.1.4) in R (version 4.4.2). After removing low quality nuclei, integrated data was normalized using *NormalizeData* function, and the top 2000 genes were used for *FindVariableFeatures* function with default parameters. *ScaleData*, *RunPCA* (npcs=30), *FindNeighbors* (ndims=30), *FindClusters* (resolution= 0.5, algorithm= 1), and *RunUMAP*(dims = 1:30) functions were used with default parameters, respectively. To remove the doublets, we used *DoubletFinder* (PCs = 1:10) with default parameters. Major cell types were annotated both based on canonical markers, including astrocytes (*GFAP*), microglia (*C3*), oligodendrocytes (*MOBP*), oligodendrocyte precursor cells (*PDGFRA*), neurons (*SLC17A7*, *GAD1*), endothelial cells (*CLDN5*), and pericytes (*PDGFRB*) (**Fig. 2**) and ROSMAP label transfer. Both methods resulted with the distribution of 36 clusters into the same cell types.

### Differential gene expression and enrichment analysis

Differentially expressed genes (DEGs) for each cluster (**Extended Data Table 5**) were identified between APOE-ε3/4, APOE-ε3/3, and APOE-ε4/4 nuclei using *FindMarkers* function of Seurat with the Wilcoxon rank-sum test, corrected with the Benjamini-Hochberg method. Cell-type-specific DEGs were analyzed separately. Gene ontology (GO), KEGG, and Reactome enrichment analyses were performed using the *clusterProfiler* and *ReactomePA* packages of R with adjusted p < 0.05 considered significant.

### Cross-cohort validation using ROSMAP data

Publicly available single-nucleus RNA-seq data from the Religious Orders Study and Memory and Aging Project (ROSMAP; https://www.synapse.org/Synapse:syn3219045) were re-analyzed to assess the reproducibility of *APOE-ε4*–associated transcriptional signatures. Raw FASTQ files were aligned to the GRCh38 reference with Cell Ranger (v6.1.2), and nuclei with fewer than 200 detected genes or more than 20% mitochondrial reads were excluded. Doublets were identified and removed using DoubletFinder (PCs 1–10). The filtered count matrices were normalized and variable features identified with Seurat (v4.1.4), followed by scaling, principal component analysis (npcs = 30), nearest-neighbor graph construction, clustering (resolution = 0.5), and UMAP embedding. For each major cell type (astrocytes, microglia, neurons, oligodendrocytes, oligodendrocyte progenitor cells, endothelial cells, and pericytes), we performed two differential expression analyses using the Wilcoxon rank-sum test with Benjamini–Hochberg correction: APOE-ε3/4 versus APOE-ε3/3 and APOE-ε4/4 versus APOE-ε3/4. Genes passing an adjusted p < 0.05 in both comparisons were retained. Finally, DEGs identified in both the NYBB and ROSMAP cohorts that exhibited consistent directionality.

### Gene prioritization scoring

Each DEG was assigned a composite priority score based on p-value (by taking -log value), average log2 fold change (absolute value of linear fold change), and expression frequency within clusters (percentage of cells expressing the gene in corresponding cell type). Genes were shortlisted based on high total score and cell-type specificity (>15% contribution from a single cluster). Genes showing consistent directionality across NYBB and ROSMAP were prioritized.

### Identification of astrocyte states corresponding to ROSMAP snSeq

Subpopulation identification of astrocyte clusters were performed as label transferring from the ROSMAP snSeq study in 424 brains^35^. Astrocyte subpopulation identities from the ROSMAP single-nucleus RNA-seq study35 (n = 424 donors) were projected onto our NYBB astrocyte nuclei using Seurat’s label-transfer workflow (v4.1.4). Briefly, both the ROSMAP reference and NYBB query assays were normalized using *SCTransform*, and the top 2,000 variable features were identified in each dataset. *FindTransferAnchors* function was run on the *SCTransform* assays (anchor.features = 2,000; dims = 1:30; k.filter = 200) to identify shared correlation structure between datasets. Anchors were filtered by L2 norm and mutual nearest neighbors, and *TransferData* was executed with weight.reduction = “*pcaproject*” and dims = 1:30 to impute ROSMAP astrocyte-state labels onto each NYBB nucleus. Predicted labels with a prediction score ≥ 0.5 were retained, and nuclei with lower confidence were marked “*unassigned*.” Transferred labels were integrated into the NYBB Seurat object and the predicted state assignments were used to stratify astrocyte nuclei into ten transcriptionally defined states (Ast.1 through Ast.10), matching the original ROSMAP nomenclature. These harmonized state annotations enabled downstream, state-resolved analyses.

### Human iPSC-derived astrocyte culture and immunostaining

Isogenic human iPSC lines expressing *APOE-ε3/ε3 or APOE-ε4/ε4* were differentiated into astrocytes as described^75^. The iPSCs were cultured under feeder-free conditions on hydrogel-coated plates using mTeSR1 medium (STEMCELL Technologies), with daily media changes. Cells were passaged upon reaching approximately 80% confluency to sustain optimal growth. Differentiation into astrocytes was performed as described^76^, and efficiency was assessed through immunostaining for astrocytic markers such as GFAP, confirming the generation of a homogeneous astrocyte population. For immunocytochemistry, astrocytes were fixed using 4% paraformaldehyde, permeabilized with 0.1% Triton X-100, and blocked with 5% normal serum to reduce non-specific binding. Cells were then incubated with primary antibodies targeting DIAPH3 (rabbit polyclonal, Proteintech, Cat #14342-1-AP, 1:300), PTPRG (rabbit polyclonal, Invitrogen, Cat #PA5-67565, 1:300), PDE4D (Rabbit polyclonal, Invitrogen, Cat #PA5-21590, 1:300) Phospho-FAK (Tyr397) (rabbit monoclonal, Invitrogen, Cat #44-625G, 1:300), Phospho-AKT (Ser473) (mouse monoclonal, Proteintech, Cat #66444-1-IG, 1:300), LPAR1 (EDG2, rabbit polyclonal, Invitrogen, Cat #PA5-105276), 1:300), and MATN2 (rabbit polyclonal, Abcam, Cat #ab238910, 1:300). Fluorescently labeled secondary antibodies conjugated to Alexa Fluor dyes (488, 555, or 647) were applied at a 1:500 dilution, and DAPI was used for nuclear staining. Immunofluorescence images were captured utilizing a Zeiss Axio Observer microscope integrated with a confocal module. Fluorescence intensities were quantified using Zeiss Zen software and subsequently normalized to DAPI for each staining.

### Proteomics analysis

Cells were collected in Eppendorf tubes and washed twice with ice-cold PBS. Cells were then lysed in a lysis buffer containing 8 M urea, 1% SDS in 0.1 M ammonium bicarbonate, and protease inhibitors (1 mini-Complete EDTA-free tablet). The lysate was cleared by centrifugation at 14,000 rpm for 30 minutes at 4 °C. The supernatant was transferred to a new tube, and the protein concentration was determined using a BCA assay (Pierce). Subsequently, 10 µg of total protein was subjected to disulfide bond reduction with 10 mM DTT (at 56 °C for 30 minutes) followed by alkylation with 10 mM iodoacetamide (at room temperature for 30 minutes in the dark). Excess iodoacetamide was quenched with 5 mM DTT (at room temperature for 15 minutes in the dark). Samples were chloroform-methanol precipitated and pellets were resuspended in 100 mM Tris-HCl (pH=8.0). Whole-cell lysates from iPSC-derived APOE-ε3/ε3 and APOE-ε4/ε4 astrocytes were processed for label-free quantitative proteomics using mass spectrometry. Protein identification and quantification were performed using MaxQuant, with differential expression determined by log2FC >1 and adjusted p < 0.05.^77^

### Immunohistochemistry on human brain tissue

Postmortem paraffin-embedded brain sections from *APOE-ε3/ε3* and *APOE-ε4/ε4* individuals were stained for relevant antibodies. Sections were imaged using confocal microscopy and analyzed using blinded quantification. Post-mortem paraffin-embedded brain sections of the BA9 prefrontal cortex from *APOE-ε3/ε3* and *APOE-ε4/ε4* individuals were obtained from the New York Brain Bank at Columbia University. The immunohistochemistry (IHC) protocol applied following the previously published procedures^3,25,65,78^. For the IHC, LAMA3 (rabbit polyclonal, Invitrogen, Cat.# PA5-52665, dilution 1:250), IGF1 (goat polyclonal, Abcam, Cat #ab106836, dilution 1:500), COL6A2 (rabbit polyclonal, Cat #PA5-65085, dilution 1:200), CDH5 (goat polyclonal, Cat #PA5-143232, dilution 1:100), Phospho-PTK2 (phospho-FAK (Tyr397) (rabbit monoclonal, Invitrogen, Cat #44-625G, 1:100)), Phospho-AKT (Ser473) (mouse monoclonal, Proteintech, Cat #66444-1-IG, 1:200), LAMA1 (mouse monoclonal, Invitrogen, Cat #MA5-31380, 1:500), LAMA2 (monoclonal mouse, Sigma, Cat #AMAB91166-100UL, 1:200), COL4 (monoclonal mouse, Invitrogen, Cat #14-9871-82, 1:100) and GFAP (Thermo Fisher, Cat #OPA1-06100, dilution 1:200) were used as primary antibodies. Secondary antibodies were goat anti-mouse Alexa Fluor 448 (Thermo Fisher, Cat.# A11001, dilution 1:500), goat anti-mouse Alexa Fluor 555 (Thermo Fisher, Cat.# A-21127, dilution 1:500), goat anti-rabbit Alexa Fluor 647 (Thermo Fisher, Cat #A32733, dilution 1:500), goat anti-chicken Alexa Fluor 488 (Thermo Fisher, Cat # A-11039, dilution 1:500), donkey anti-mouse Alexa Fluor 488 (Thermo Fisher, Cat #A32766, dilution 1:500), donkey anti-goat Alexa Fluor 555 (Thermo Fisher, Cat #A32816, dilution 1:500), and donkey anti-rabbit Alexa Fluor 647 (Thermo Fisher, Cat #A32795, dilution 1:500).

### Immunohistochemistry on mouse brain tissue

Brains from targeted *APOE* replacement mouse models^79–81^ were stained for Phospho-PTK2 (phospho-FAK (Tyr397) (rabbit monoclonal, Invitrogen, Cat #44-625G, 1:100)), Phospho-AKT (Ser473) (mouse monoclonal, Proteintech, Cat #66444-1-IG, 1:200), and GFAP (Thermo Fisher, Cat # OPA1-06100, dilution 1:200). Goat anti-mouse Alexa Fluor 555 (Thermo Fisher, Cat #A-21127, dilution 1:500), goat anti-rabbit Alexa Fluor 647 (Thermo Fisher, Cat #A32733, dilution 1:500), goat anti-chicken Alexa Fluor 488 (Thermo Fisher, Cat # A-11039, dilution 1:500) were used as secondary antibodies as previously described^65^.

### Confocal Imaging

Immunostained human and mouse brain sections were mounted in Fluoromount-G and imaged on a Zeiss LSM800 laser-scanning confocal microscope equipped with 405 nm, 488 nm, 555 nm, and 639 nm diode lasers. A Plan-Apochromat 10x/20x/40x objectives were used throughout. To minimize channel crosstalk, sequential line scanning (multi-track mode) was employed, with laser power and detector gain optimized on negative controls and held constant across all samples. Pinhole size was set to 1 Airy unit for each channel, yielding a default optical section thickness. Images were acquired at 12-bit depth and a pixel size of ∼0.13 µm (1024 × 1024 frame size). Raw .czi files were exported to Zeiss Zen Blue (v3.2) for initial processing. All quantifications were performed on individual optical sections to avoid projection artifacts. Analyses were performed blind to genotype, sampling five nonoverlapping fields per section and three sections per animal or human donor. Aggregate statistics were exported for downstream statistical testing.

### Protein-protein interaction analysis

Prioritized proteins (DIAPH3, PTPRG, PDE4D, LPAR1, MATN2) were queried in STRING v12 (https://string-db.org) using the Homo sapiens database. Interactions from “Experiments,” “Databases,” “Co-expression,” and “Neighborhood” evidence channels were included, with a medium confidence score cutoff of 0.4 to balance sensitivity and specificity. The resulting network, comprising direct (first-shell) and indirect (second-shell) interactors, was exported as a TSV file and imported into Cytoscape (v3.9.1). Using the NetworkAnalyzer tool, we computed topological metrics including degree centrality, betweenness centrality, and clustering coefficient for each node. To identify densely interconnected modules, we applied the MCODE plugin (v2.0.0) with default parameters (degree cutoff = 2, node score cutoff = 0.2, k-core = 2, max depth = 100), highlighting subclusters of potential functional relevance. Hub proteins were defined as nodes in the top 10% of degree centrality, and their functional annotations were inspected manually to contextualize their role in focal adhesion and cytoskeletal remodeling.

### Statistical analysis

All quantitative data are presented as mean ± s.e.m. Statistical tests were chosen based on data distribution and experimental design, with normality assessed by the Shapiro–Wilk test and homogeneity of variances by Levene’s test. For two-group comparisons, we used two-tailed Student’s t-tests when both normality and equal variance criteria were met, and Mann–Whitney U tests otherwise. For comparisons among three or more groups, we applied one-way ANOVA followed by Tukey’s post hoc test; if data violated ANOVA assumptions, we used the Kruskal–Wallis test with Dunn’s *post hoc* correction. In experiments with two independent variables, two-way ANOVA with Sidak’s multiple comparisons correction was performed. Single-nucleus and single-cell differential expression analyses were carried out in Seurat (v4.1.4): we used the Wilcoxon rank-sum test for cell-level comparisons and incorporated donor identity as a random effect when appropriate. Statistical power for detecting cell-type-specific expression changes was estimated a priori using the snPower package, targeting ≥80% power to detect a log₂ fold change of 0.25 at α = 0.05. Multiple testing correction across genome-wide and pathway-level analyses employed the Benjamini–Hochberg procedure, with FDR < 0.05 considered significant. Missing values were handled by pairwise exclusion; no imputation was performed. All assays included at least three independent biological replicates unless otherwise stated, and exact P-values are provided in figures.

## Data Availability

Raw snRNA-seq and snATAC-seq data are deposited in synapse.org. Proteomics datasets are deposited in ProteomeXchange PRIDE. Accessions available upon reasonable request.

---

**Extended Data Figure 1 Top 20 enriched pathways based on differentially expressed genes in *APOE-ε4* astrocytes.** Chord diagrams visualize the top 20 enriched terms from four functional categories based on differentially expressed genes (DEGs) identified in *APOE-ε4/4* versus *APOE-ε3/3* astrocytes. The diagrams display the connections between individual DEGs and their associated terms for Gene Ontology (GO) Biological Process (top left), GO Molecular Function (top right), GO Cellular Component (bottom left), and KEGG Pathway (bottom right). Notable biological processes include extracellular matrix organization, cell adhesion, and system development. In the molecular function category, enriched terms include integrin binding, cytokine activity, and peptidase regulation. Cellular component analysis highlights associations with the plasma membrane, basement membrane, and extracellular matrix structures. KEGG pathway enrichment reveals significant overlap with focal adhesion, ECM–receptor interaction, PI3K-AKT signaling, and Alzheimer’s disease pathways. These visualizations highlight the broad functional impact of *APOE-ε4* on astrocytic signaling, adhesion, and structural organization.

**Extended Data Figure 2** *APOE-ε4* astrocytes display altered expression of genes involved in multiple signaling pathways related to adhesion and survival. Heatmaps show log2 fold changes of differentially expressed genes (DEGs) across astrocyte clusters in *APOE-ε4/4* versus *APOE-ε3/3* brains for four major signaling pathways: Adherens junctions, Insulin signaling, Neurotrophin signaling, and MAPK signaling. Red and blue represent upregulation and downregulation, respectively, with color intensity corresponding to fold change magnitude. The adherens junction pathway includes structural components such as *CTNNA1, ACTN2,* and *PTPRU*, indicating dysregulated cell-cell contact. Within the insulin signaling pathway, alterations in key regulators including *AKT1, IRS2, PIK3R1,* and *IGF1R* suggest *APOE-ε4*-associated disruptions in survival and metabolic signaling. Neurotrophin signaling changes involve genes such as *MAPK1, CAMK2A*, and *NTRK2*, further supporting impaired trophic support in *APOE-ε4* astrocytes. MAPK signaling, shown in the rightmost panel, displays broad transcriptional changes, including in *MAP2K1*, *JUN, DUSP1,* and *RAF1*, which are key mediators of stress and inflammatory signaling. These data collectively illustrate that *APOE-ε4* astrocytes undergo widespread reprogramming of intracellular signaling networks that converge on cytoskeletal remodeling, metabolic function, and adhesion stability.

**Extended Data Figure 3 Correlation matrix of gene-level metrics used in prioritization scoring.** Pearson correlation analysis was performed to assess the relationship between six gene-level variables used in prioritizing differentially expressed genes across astrocyte clusters: p-value, log2 fold change (logFC), adjusted p-value (p-adj), average expression (AveExp), aggregated expression (AggExp), and percentage of expressing cells (%Exp). The lower triangle displays Pearson correlation coefficients as a color scale (blue to orange), while the upper triangle shows exact correlation values with asterisks indicating statistically significant correlations (p < 0.05). Strong positive correlations were observed among AveExp, AggExp, and %Exp, consistent with these metrics representing gene abundance and detection frequency across cells. In contrast, p-values and p-adj values showed weak or no correlation with expression-based metrics, supporting their independent contribution to the prioritization score. These findings justify the use of multiple uncorrelated parameters in the gene scoring framework.

**Extended Data Figure 4 Seurat label transfer and subcluster-resolved expression of prioritized focal adhesion genes in NYBB astrocytes. (a)** Schematic of the label-transfer workflow, in which annotated astrocyte states from the ROSMAP reference^35^ were projected onto the NYBB snRNA-seq astrocyte dataset using a nearest-neighbor machine-learning algorithm. **(b)** UMAP embedding of NYBB astrocytes colored by the nine “Ast” states inferred from ROSMAP. Each dot is one nucleus; labels indicate subcluster identity. **(c)** Heat map of astrocyte state markers in NYBB snSeq after label transfer from ROSMAP snSeq. **(d)** Feature plots of DIAPH3, MATN2, PDE4D, PTPRG, and LPAR1 expression (log-normalized counts) over the same UMAP projection. Color intensity scales with expression level (see legend at right of each panel). **(e)** Violin + dot plots showing the distribution of expression levels for each gene across the ten Astrocyte subclusters (Ast-110). Black dots are individual nuclei; violins indicate kernel density.

**Extended Data Figure 5 Prioritized gene expression is independent of AD status. (a)** Violin plots showing normalized expression levels of prioritized focal adhesion-related genes (*PDE4D, DIAPH3, PTPRG, and LPAR1*) in astrocytes across four groups: APOE-ε3/ε3 with AD (e3-e3_AD), APOE-ε3/ε3 without AD (e3-e3_NonAD), APOE-ε3/ε4 with AD (e3-e4_AD), and APOE-ε3/ε4 without AD (e3-e4_NonAD). Expression patterns are consistent between AD and non-AD individuals within the same genotype group. **(b)** Combined violin plots comparing expression of the same four genes across all groups in astrocytes, colored by genotype and AD status.

**Extended Data Figure 6 Cell-type and *APOE*-genotype-specific chromatin accessibility at prioritized genes.** Genome browser tracks showing aggregate snATAC-seq signal (normalized Tn5 insertion density) across major brain cell types (astrocytes, endothelial cells, excitatory neurons, inhibitory neurons, oligodendrocytes, OPCs) at the DIAPH3, LPAR1, PTPRG, PDE4D, and MATN2 loci (left column). Each colored track (left panels) corresponds to one cell type. The panels on the right show the log₂ accessibility distribution for APOE-ε3/3 (red) versus APOE-ε3/4 (blue) nuclei. Below each browser view are called peak regions (red tick marks) and the corresponding gene model (exons, UTRs).

**Extended Data Table 1:** Demographics of the cohort indicating age, sex, APOE genotype, Braak stage, clinical diagnosis, cerebral amyloid angiopathy (0: none reported, 1: mild, 2: moderate, 3: severe), and arteriolosclerosis (0: none reported, 1: mild, 2: moderate, 3: severe).

**Extended Data Table 2:** Nuclei numbers in APOE genotypes and clinical AD status NYBB snSeq

**Extended Data Table 3:** Nuclei numbers in APOE genotypes and cell types in NYBB snSeq

**Extended Data Table 4:** Proportion of cells according to APOE genotypes in NYBB snSeq

**Extended Data Table 5:** NYBB snSeq DEGs in APOE3/4 versus APOE3/3

**Extended Data Table 6:** ROSMAP snSeq DEGs APOE3/4 vs APOE3/3

**Extended Data Table 7:** Common 319 genes between NYBB-scSeq and ROSMAP

**Extended Data Table 8:** Priority scores for all clusters and 319 common genes

**Extended Data Table 9:** ROSMAP snSeq DEGs APOE4/4 vs APOE3/4

**Extended Data Table 10:** NYBB snSeq DEGs APOE4/4 vs APOE3/4

**Extended Data Table 11:** Isogenic iPSC-derived human astrocyte global proteomics results

## Acknowledgements

This work was supported by National Institute on Aging R01 AG067501 (Genetic Epidemiology and Multi-Omics Analyses in Familial and Sporadic Alzheimer’s Disease Among Secular Caribbean Hispanics and Religious Order) (RM, BNV, CK) and National Institute on Aging RF1 AG066107 Epidemiological Integration of Genetic Variants and Metabolomics Profiles in Washington Heights Columbia Aging Project (RM, BNV, CK), Taub Institute Grants for Emerging Research (TIGER) (CK), Thompson Family Foundation Program for Accelerated Medicine Exploration in Alzheimer’s Disease and Related Disorders of the Nervous System (TAME-AD) (CK), Carol and Gene Ludwig Family Foundation (CK, AJL, BNV), Toffler Scholar Program (PB), and P30 AG066462 Alzheimer’s Disease Research Center (AFT). Shared Resource of the Herbert Irving Comprehensive Cancer Center (HICCC) was supported from the P30 Cancer Center Support Grant. This research was funded in part through the NIH/NCI Cancer Center Support Grant P30CA013696 and used the Genomics and High Throughput Screening Shared Resource. The content of this publication is solely the responsibility of the authors and does not necessarily represent the official views of the National Institutes of Health. We would like to thank New York Brain Bank for post-mortem human brain sections and Taub Institute Imaging Platform; the contributors, who collected samples used in this study; the patients and families for their participation, without whom these studies would not have been possible; Erin Bush (Single Cell Analysis Core and Columbia Genome Center, Sulzberger Genome Center) for single nucleus sequencing, Dr. Rajesh Soni (HICC Proteomics and Structural Biology Facility) for proteomics, and Dr. Tal Nuriel for mouse brains used in this study.

